# A simple model to assess Wuhan lock-down effect and region efforts during COVID-19 epidemic in China Mainland

**DOI:** 10.1101/2020.02.29.20029561

**Authors:** Zheming Yuan, Yi Xiao, Zhijun Dai, Jianjun Huang, Yuan Chen

## Abstract

Since COVID-19 emerged in early December, 2019 in Wuhan and swept across China Mainland, a series of large-scale public health interventions, especially Wuhan lock-down combined with nationwide traffic restrictions and Stay At Home Movement, have been taken by the government to control the epidemic. Based on Baidu Migration data and the confirmed cases data, we identified two key factors affecting the later (*e*.*g* February 27, 2020) cumulative confirmed cases in non-Wuhan region (*y*). One is the sum travelers from Wuhan during January 20 to January 26 (*x*_1_), which had higher infected probability but lower transmission ability because the human-to-human transmission risk of COVID-19 was confirmed and announced on January 20. The other is the “seed cases” from Wuhan before January 19, which had higher transmission ability and could be represented with the confirmed cases before January 29 (*x*_2_) due to a mean 10-day delay between infection and detection. A simple yet effective regression model then was established as follow: *y*= 70.0916+0.0054×*x*_1_+2.3455×*x*_2_ (*n* = 44, *R*^2^ = 0.9330, *P*<10^−7^). Even the lock-down date only delay or in advance 3 days, the estimated confirmed cases by February 27 in non-Wuhan region will increase 35.21% or reduce 30.74% - 48.59%. Although the above interventions greatly reduced the human mobility, Wuhan lock-down combined with nationwide traffic restrictions and Stay At Home Movement do have a determining effect on the ongoing spread of COVID-19 across China Mainland. The strategy adopted by China has changed the fast-rising curve of newly diagnosed cases, the international community should learn from lessons of Wuhan and experience from China. Efforts of 29 Provinces and 44 prefecture-level cities against COVID-19 were also assessed preliminarily according to the interpretive model. Big data has played and will continue playing an important role in public health.

## 1 Introduction

Since Corona Virus Disease 2019 (COVID-19) caused by SARS-CoV-2, a novel coronavirus, emerged on December 1, 2019 in Wuhan City, Hubei Province, China (Huang *et al*, 2020), the overall confirmed cases in China had reached 78,959 by the end of February 27, 2020, and a total of 2,791 people had died of the disease. COVID-19 had also spread to other 50 country with the confirmed cases and deaths were 4,696 and 67 respectively, by the end of February 27, 2020. The World Health Organization declared the COVID-19 epidemic as an international public health emergency on January 30, 2020.

To prevent further dissemination of SARS-CoV-2, 31 Provinces in China Mainland had raised their public health response level to the highest state of emergency (level-1) by January 29, 2020. The Chinese government has implemented a series of large-scale public health interventions to control the epidemic, many of which have far exceeded what International Health Regulations required, especially Wuhan lock-down, nationwide traffic restrictions and Stay At Home Movement. Wuhan had prohibited all transport in and out of the city as of 10:00 on January 23, 2020, this is maybe the largest quarantine/movement restriction in human history to prevent infectious disease spread (Tian *et al*, 2020). Hundreds of millions Chinese residents, including 9 million Wuhan residents, have to reduce even stop their inter-city travel and intra-city activities due to these strict measures. Due to Wuhan lock-down, Kucharski *et al* (2020) estimated that the median daily reproduction number had declined from 2.35 of January 16 to 1.05 of January 30, Tian *et al*. (2020) estimated that the dispersal of infection to other cities was deferred 2.91 days (CI: 2.54-3.29). However, Read *et al* (2020) suggested that travel restrictions from and to Wuhan city are unlikely to be effective in halting transmission across China; with a 99% effective reduction in travel, the size of the epidemic outside of Wuhan may only be reduced by 24.9% on February 4. Do these large-scale public health interventions really work well in China? Besides, hundreds of officials were fired or appointed rapidly according to their incompetent or outstanding performances during the epidemic. How to assess the efforts of different regions in China Mainland against COVID-19?

Here we present a simple yet effective model based on Baidu Migration data and the confirmed cases data to quantify the consequences and importance of Wuhan lock-down combined with nationwide traffic restrictions and Stay At Home Movement on the ongoing spread of COVID-19 across China Mainland, and preliminarily assess the efforts of 29 Provinces and 44 prefecture-level cities during the epidemic.

## 2 Results

### 2.1 Effect of COVID-19 on human mobility of China Mainland residents

Due to Wuhan lock-down, more than 9 million residents were isolated in Wuhan City since January 23, 2020, only 1.2 million travelers from or to Wuhan during January 24 to February 15, 2020 according to Baidu Migration. The travelers were down 91.61% and 91.62%, compared to the same period last year (14 million) and the first 23d (14 million from January 1 to January 23, 2020), respectively (Fig.1).

**Fig. 1.**
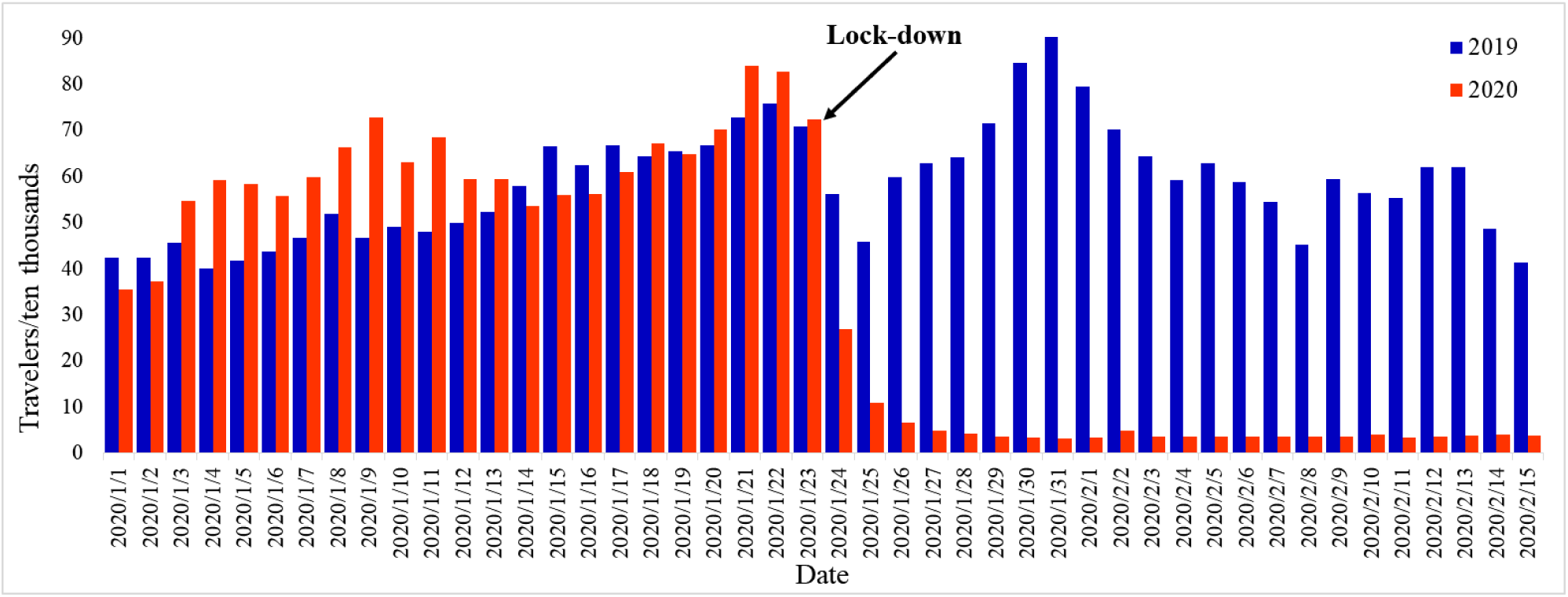
Number of travelers from or to Wuhan during January 1 to February 15, 2020 and the same period last year

Due to nationwide traffic restrictions, only 185 million travelers left 316 cities during January 24 to February 15, 2020 according to Baidu Migration. The emigrants were down 69.77% and 67.60%, compared to the same period last year (611 million) and the first 23d (570 million), respectively (Fig.2).

**Fig.2.**
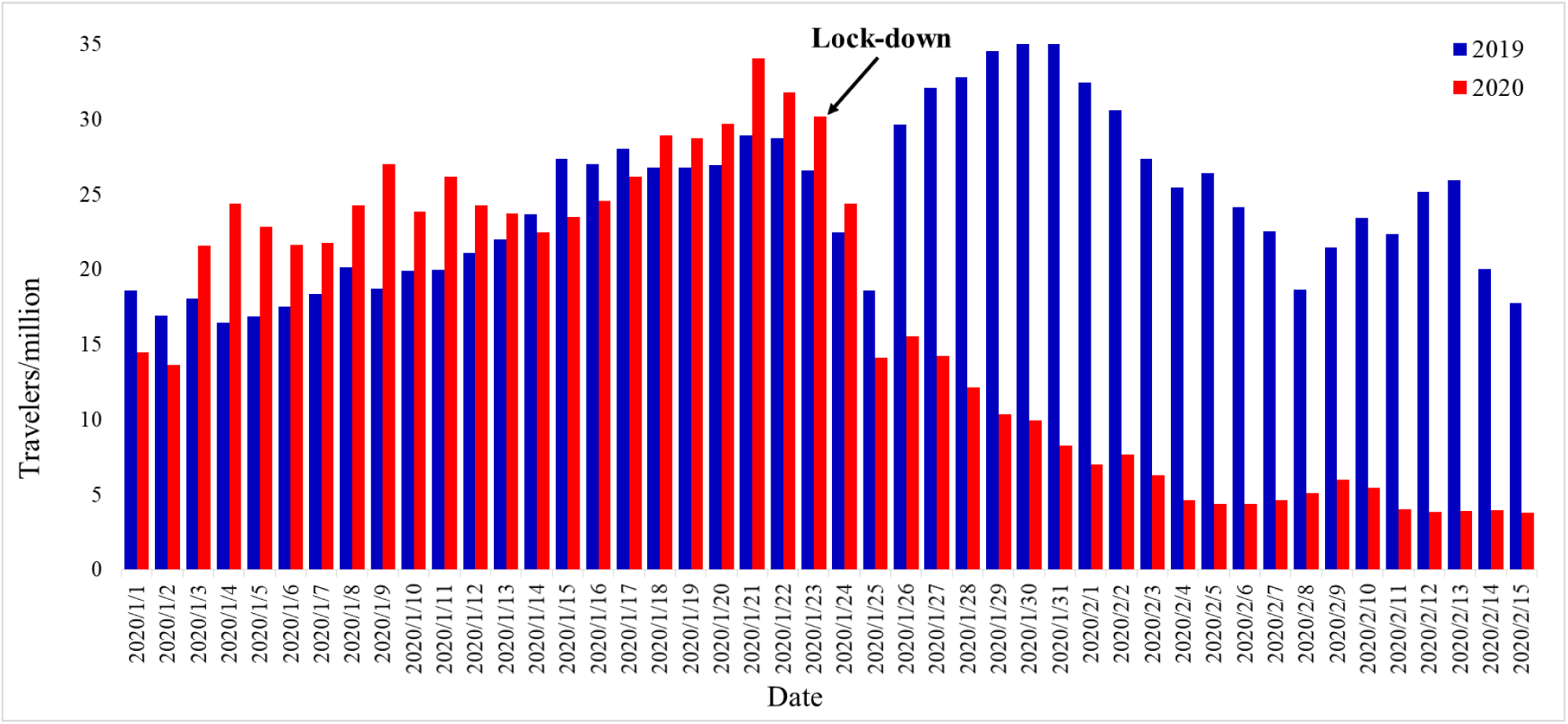
Number of travelers left 316 cities during January 1 to February 15, 2020 and the same period last year

Due to Stay At Home Movement, the mean intensity of intra-city activities for 316 cities was 2.61/d during January 24 to February 15, 2020 according to Baidu Migration. It was down 42.42% and 50.27% when compared to the same period last year (4.53/d) and the first 23d (5.25/d), respectively (Fig.3).

**Fig.3.**
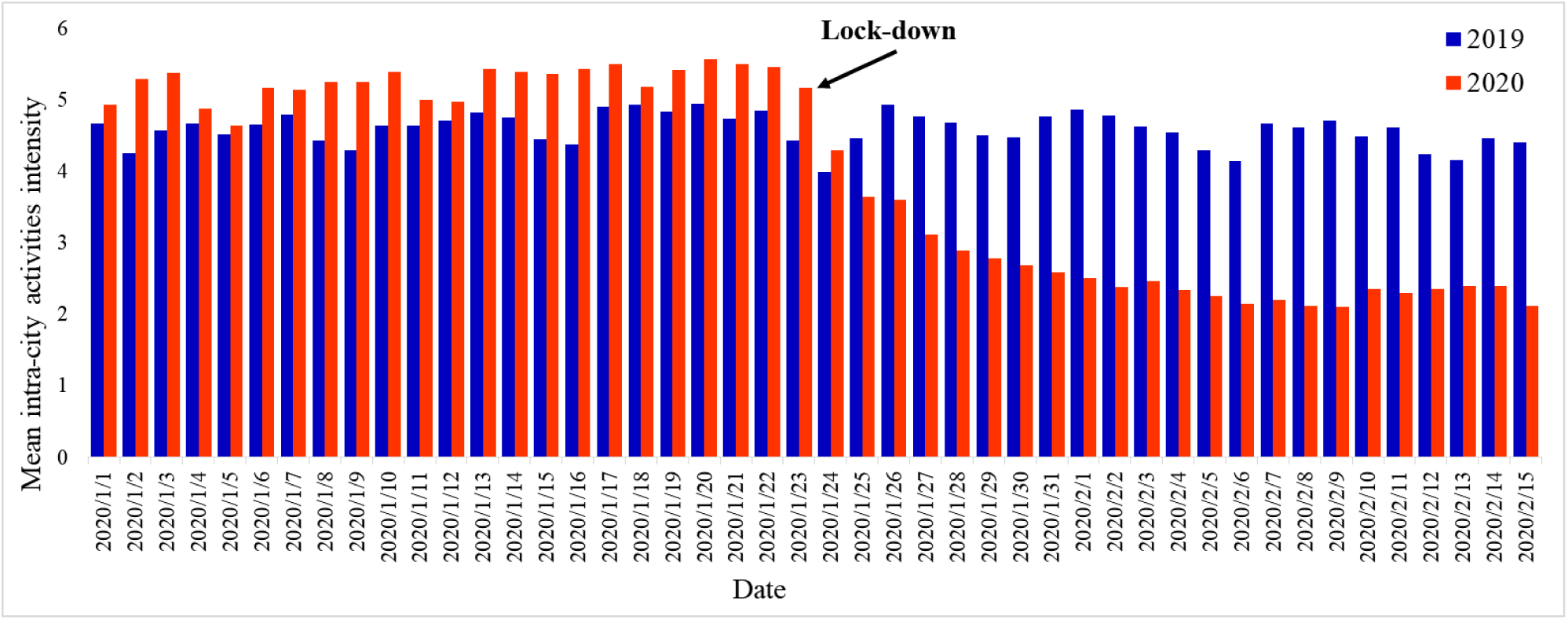
The mean intensity of intra-city activities for 316 cities during January 1 to February 15, 2020 and the same period last year

Obviously, COVID-19 greatly reduced the human mobility of China Mainland.

### 2.2 Two key factors affecting the later cumulative confirmed cases of each non-Wuhan region

We consider 44 regions in China Mainland which accept travelers from Wuhan, including 29 Provinces (Tibet was excluded since only one confirmed case was reported) and 15 prefecture-level cities in Hubei province. We noticed that the number of confirmed cases between non-Hubei and Hubei were closer in the early period. For example, the number of cumulative confirmed cases by the end of January 26 in Chongqing (non-Hubei) and Xiaogan (Hubei) were 110 and 100, respectively. Their cumulative confirmed cases by the end of February 27, however, were 576 and 3,517, respectively. We surmise that this is partly because Xiaogan has received more infected cases from Wuhan than Chongqing since the human-to-human transmission risk of COVID-19 was confirmed and announced on January 20.

This surmise was confirmed by Fig.4. The proportion of travelers from Wuhan accepted by Hubei regions to the total travelers from Wuhan increased rapidly from 70% (before January 19) to 74% (January 20), even over 77% after January 26. So we concluded that the first key factor affecting the later (*e*.*g*. February 27) cumulative confirmed cases of each non-Wuhan region is the sum immigrants from Wuhan during January 20 to January 26 (few immigrants after January 27). These immigrants had higher infected probability but lower transmission ability because the susceptible strengthened protection awareness and measures after the declaration of human-to-human transmission.

**Fig.4.**
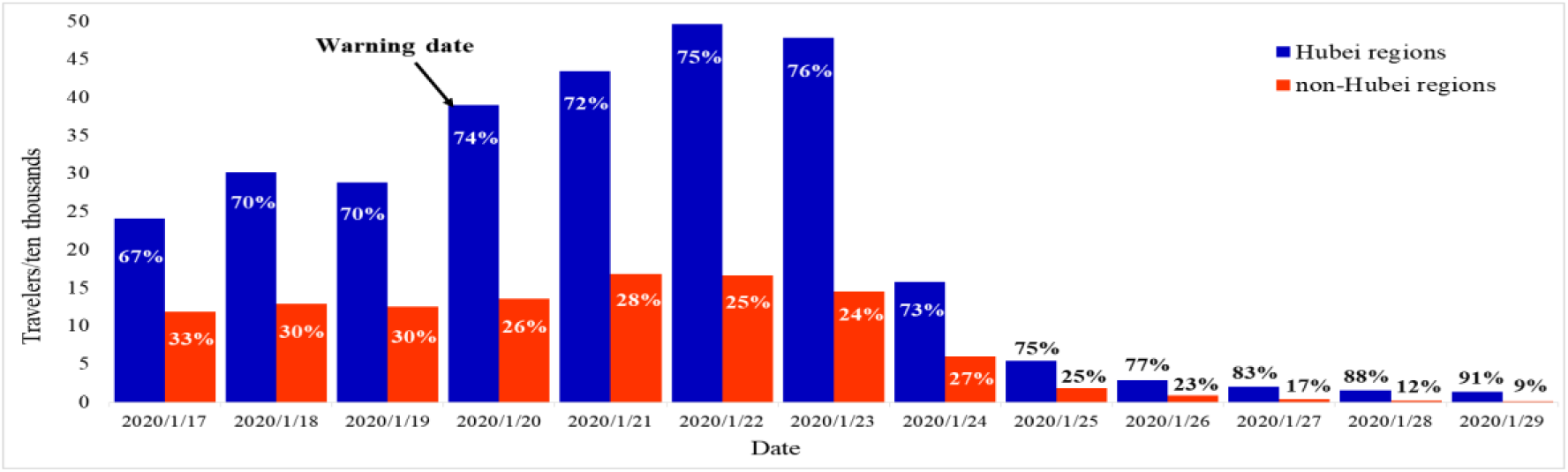
Size and proportion of travelers from Wuhan accepted by Hubei regions and non-Hubei regions before and after warning date

The second key factor is the sum number of the infected immigrants from Wuhan before January 19. According to the recent report, there is a mean 10-day delay between infection and detection, comprising a mean about 5 day incubation period and a mean 5 day delay from symptom onset to detection of a case (Imai *et al*, 2020; Yang *et al*, 2020; Li *et al*, 2020). So the second key factor can be represented with the number of cumulative confirmed cases by the end of January 29. These “seed cases” had higher transmission ability because the susceptible had no any protection measure. A simple regression model was established as follow.

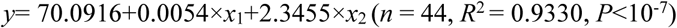

Here, *y* is the number of cumulative confirmed cases by February 27 of each non-Wuhan region, *x*_1_ is the sum number of immigrants from Wuhan during January 20 to January 26 of each non-Wuhan region, and *x*_2_ is the number of cumulative confirmed cases by January 29 of each non-Wuhan region.

The standard regression coefficients of *x*_1_ and *x*_2_ are 0.6620 and 0.3796 respectively, indicating that *x*_1_ is more important than *x*_2_ for determining *y*. The observed and expected values of the cumulative confirmed cases by February 27 of each non-Wuhan regions were shown in Fig.5.

**Fig.5.**
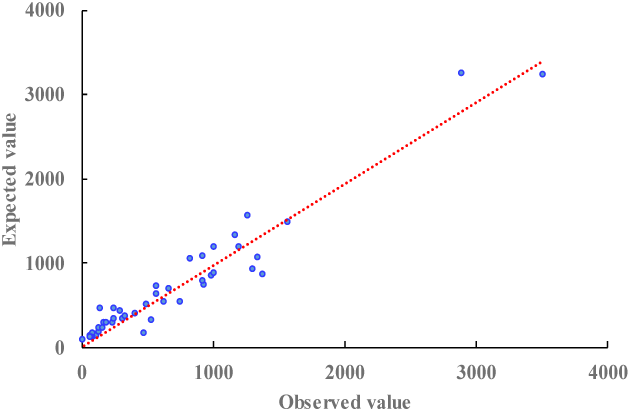
The observed and expected values of the cumulative confirmed cases by February 27 of each non-Wuhan region

### 2.3 Effect of Wuhan lock-down combined with nationwide traffic restrictions and Stay At Home Movement on the ongoing spread of COVID-19 across China Mainland

To evaluate the effect of Wuhan lock-down, *x*_2_, the number of cumulative confirmed cases by January 29, should be fixed. Then we assumed three different lock-down plans besides the real plan (Table 1).

**Table 1.**
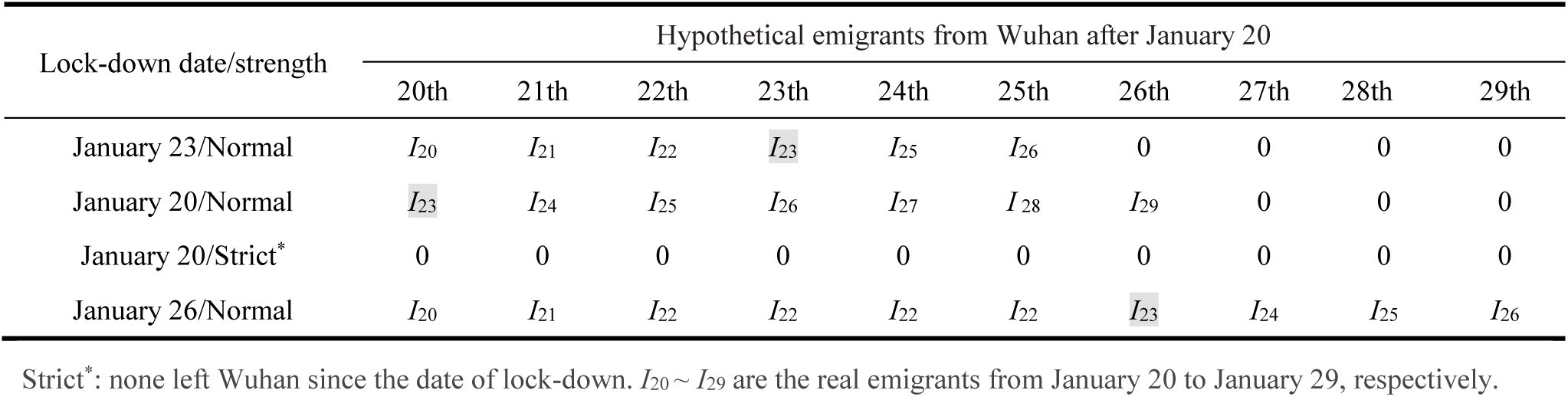
Different lock-down plans and the hypothetical emigrants from Wuhan after January 20

According to Table 1, even the lock-down date only delay 3 days, the estimated sum number of cumulative confirmed cases by February 27 in 44 non-Wuhan region will increase 35.21% (41,477 cases), compared with the real plan (30,675 cases). On the contrary, even the lock-down date only in advance 3 days with normal and strict strength, the estimated number will reduce 30.74% (21,245 cases) and 48.59% (15,768 cases), respectively. It is clearly that Wuhan lock-down combined with nationwide traffic restrictions and Stay At Home Movement have a determining effect on the ongoing spread of COVID-19 across China Mainland according to our interpretative model.

### 2.4 Preliminarily assess region efforts against COVID-19

It’s not fair to assess the effort of different regions only based on the final number of cumulative confirmed cases due to the difference of the number of immigrants from Wuhan. The aforementioned interpretative model has taken this difference into account and the results are listed with 5 grade (Excellent, Good, Normal, Poor, Very poor) according to standard residuals (Table 2 and Table 3). We emphasize that this is only a preliminary evaluation and the results are for reference only.

**Table 2.** Preliminarily assess 29 Provinces efforts against COVID-19

| Province | Obs.* | Exp. | Standard residuals | Grade | Province | Obs.* | Exp. | Standard residuals | Grade |
| --- | --- | --- | --- | --- | --- | --- | --- | --- | --- |
| Guizhou | 146 | 454 | -1.63 | Excellent | Tianjin | 136 | 178 | -0.22 | Normal |
| Henan | 1272 | 1553 | -1.49 | Excellent | Jilin | 93 | 125 | -0.17 | Normal |
| Hunan | 1017 | 1189 | -0.91 | Good | Shanghai | 337 | 359 | -0.12 | Normal |
| Fujian | 296 | 423 | -0.67 | Good | Liaoning | 121 | 133 | -0.07 | Normal |
| Yunnan | 174 | 295 | -0.64 | Good | Hebei | 318 | 328 | -0.05 | Normal |
| Shanxi | 133 | 225 | -0.49 | Normal | Zhejiang | 1205 | 1193 | 0.06 | Normal |
| Guangxi | 252 | 341 | -0.47 | Normal | Beijing | 410 | 394 | 0.09 | Normal |
| Gansu | 91 | 170 | -0.42 | Normal | Jiangsu | 631 | 535 | 0.51 | Poor |
| Qinghai | 18 | 89 | -0.37 | Normal | Anhui | 990 | 846 | 0.76 | Poor |
| Hainan | 168 | 232 | -0.34 | Normal | Jiangxi | 935 | 732 | 1.08 | Very poor |
| Neimenggu | 75 | 131 | -0.30 | Normal | Sichuan | 538 | 323 | 1.14 | Very poor |
| Shaanxi | 245 | 294 | -0.26 | Normal | Shandong | 756 | 539 | 1.15 | Very poor |
| Chongqing | 576 | 622 | -0.24 | Normal | Guangdong | 1348 | 1061 | 1.52 | Very poor |
| Ningxia | 72 | 116 | -0.23 | Normal | Heilongjiang | 480 | 165 | 1.67 | Very poor |
| Xinjiang | 76 | 119 | -0.23 | Normal |  |  |  |  |  |

\*: the confirmed cases by February 27.

**Table 3.**
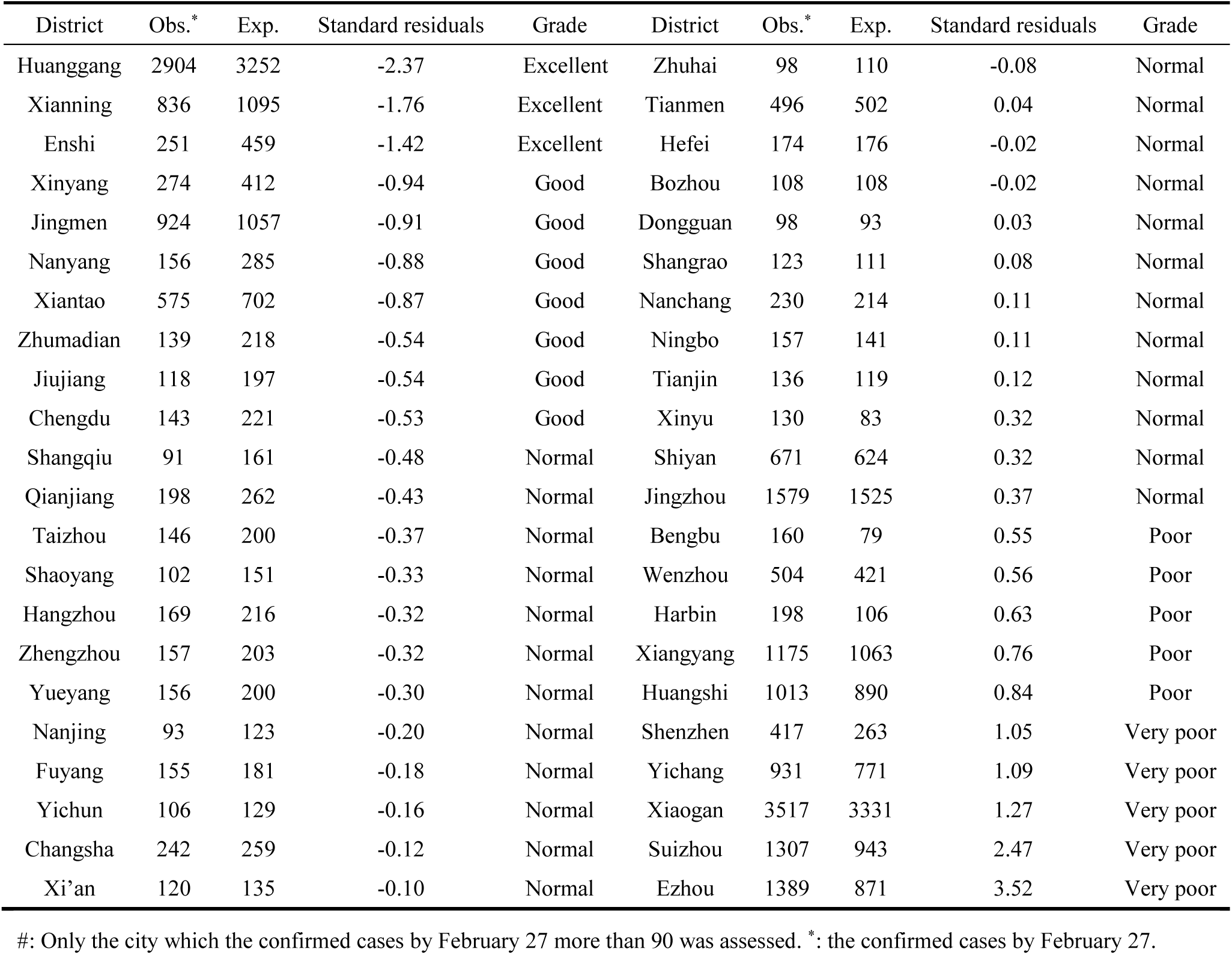
Preliminarily assess 44 prefecture-level cities efforts against COVID-19^#^

## 3 Dataset and Methods

The human mobility data on inter-city travel and intra-city activity from January 1, 2020 to February 21, 2020 (including the same period data in 2019) in China Mainland is from Baidu Migration (http://qianxi.baidu.com). The inter-city travel population of each city is represented with immigration and emigration index, the travelers proportions of different destination and departure, on the level of Province and city (but only the top 100 cities), are also listed. The intra-city activity intensity of each city is represented with the index (not proportion) of activity population to total population.

Real immigration and emigration populations of Beijing and Shanghai, emigration populations of Wuhan, Nanjing, Qingdao, Shenzhen and Foshan during the 2019 Spring Festival travel rush (40d) are from their Official website of the Transportation Commission. We estimate one Baidu Migration index is about equal to 56,137 travelers (Fig.6).

**Fig.6.**
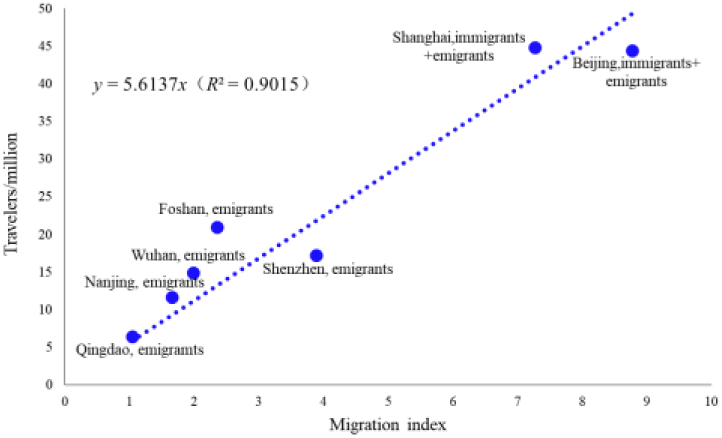
Relationship between Baidu Migration index and the number of real travelers

The confirmed case data on each Province and prefecture-level city are from the National Health Commission of China (http://www.nhc.gov.cn/xcs/yqtb/list_gzbd.shtml) and its affiliates.

## 4 Discussion

### 4.1 Accuracy of estimating human mobility based on Baidu Migration

Baidu Migration recorded more types of spatial displacement, including airplane, high-speed rail, ship, coach and private car, so it theoretically has higher accuracy. The real inter-city travel population of each city, however, is difficult to estimate because Baidu Migration can only list the migration index and the proportion. Based on a complicated hyper-network model (Chan and Hsu, 2015), Zhang and Xiu (2020, unpublished, https://blog.csdn.net/BigDataDigest/article/details/104197152) estimated that the emigration population of Shenzhen and the immigration population of Beijing from January 10 to January 23, 2020 (14d) were 7,469,192 and 3,570,184, and the corresponding Baidu Migration index were 245.48 and 116.95. One Baidu Migration index is about equal to 30,427-30,526 travelers according to their model, this is far below our estimated value 56,137. The sum immigration index of Wuhan from the first day of the Spring Festival travel rush (January 10) to the last day before lock-down (January 22) is 95.98, then the two model estimated travelers are 2.9 million and 5.4 million, respectively. News Release Conference of Wuhan on 26 January confirmed that more than 5 million people had left Wuhan after 10 January due to the Spring Festival travel rush and epidemic. Our estimated value is closer to official reports.

It is worth noting that the Baidu Migration index still has the following disadvantages for estimating real traveler population. 1) The mobility behavior of a large number of groups un-connected to Baidu Map and third-party users has not been recorded. 2) The spatial displacement of users is recorded only within 8 hours. 3) Most of the trips are disassembled and not fully identified. For example, one user travel from A, pass through B and arrive final destination C, and the user also has location information in C by coincidence, then this trip will be disassembled into A-B and B-C. Regardless, big data has and will continue to play an important role in public health.

### 4.2 How many Wuhan residents were infected with SARS-CoV-2

Several models have estimated the number of infected individuals in Wuhan. Based on the domestic and international confirmed case, Imai *et al* (2020) estimated that the total number of infected individuals was 21,022 (CI: 11,090-33,490) by January 22. Based on the number of cases exported from Wuhan internationally, number of international flights arriving in Wuhan and the latest human mobility data from Tencent, Wu *et al* (2020) estimated that the total number was 75,815 (CI: 37,304-130,330) by January 25. Based on the meta-data of five countries’ evacuation action from January 29 to February 2, Zhao *et al* (2020) estimated that almost 110,000 (CI: 40,000-310,000) individuals were infected with SARS-CoV-2 in Wuhan by February 2. Wang *et al* (2020) estimated 4 phases divided by the dates when various levels of prevention and control measures were taken in effect in Wuhan, the number of infections would reach the peak 58,077-84,520 or 55,869-81,393 in late February.

There are 9.48 million residents in Wuhan around January 26, the cumulative confirmed cases are 2,261 by January 29. We estimate at least 56,916 people were infected in Wuhan according to our model (up to February 27 the confirmed cases are 48,137). In other words, more than 8,000 undetected but infected individuals still wait for checking out in the center of epidemic storm. Wuhan still has much to do.

### 4.3 Lesson from Wuhan and experience from China

SARS-CoV-2 has diversity transmission approaches, including respiratory (mouth foam) and contact routes which have been confirmed, as well as aerosol and digestive (fecal-oral) routes which cannot be ruled out (National Health Commission of China, 2020). It is also highly concealed according to the recent report of transmission of the virus from asymptomatic and mild individuals (Zhang *et al*, 2020; Sanche *et al*, 2020). Initially, the basic reproductive number, *R*_0_, was estimated to be 2.2 (CI: 1.4-3.9) by Li *et al* (2020), 2.68 (CI: 2.47-2.86) by Wu *et al* (2020), 3.6-4.0 by Imai *et al* (2020) and Read *et al* (2020), and 3.77 (range 2.23-4.82) by Yang *et al* (2020), respectively. At the beginning of the outbreak, the infected individuals may be greatly underestimated due to the asymptomatic transmissions, insufficient sensitivity of diagnostic reagents and delayed diagnosis. The latest estimated value of *R*_0_ and the control reproductive number is 4.7-6.6 (Steven *et al*, 2020) and 6.47 (CI: 5.71-7.23) (Tang *et al*, 2020), respectively.

SARS-CoV-2 is highly contagious, Wang *et al* (2020) projected that without any control measure the infected population would exceed 200,000 in Wuhan by the end of February. As for Steven *et al* (2020) and Read *et al* (2020), the estimated number was 233,400 (CI: 38,757-778,278) by the end of January and 191,529 (CI: 132,751-273,649) by February 4, respectively. Up to February 27, the confirmed cases are 48,137.

The only lesson that humans have learned from history is that humans have not learned anything from history. Clearly, Wuhan has not learned anything from the SARS epidemic in 2003, now she is suffering from her early delays. Fortunately, China Government has implemented a series of large-scale public health interventions to control the epidemic. In fact, many prevention and control measures taken by China, especially Wuhan lock-down, nationwide traffic restrictions and Stay At Home Movement, go far beyond the requirements for responding to emergencies, setting new benchmarks for epidemic prevention in other countries. The Chinese method has proven to be successful. The strategy adopted by China has changed the fast-rising curve of newly diagnosed cases, and the simplest and most direct thing that can explain this is the data (Fig.7).

**Fig.7.**
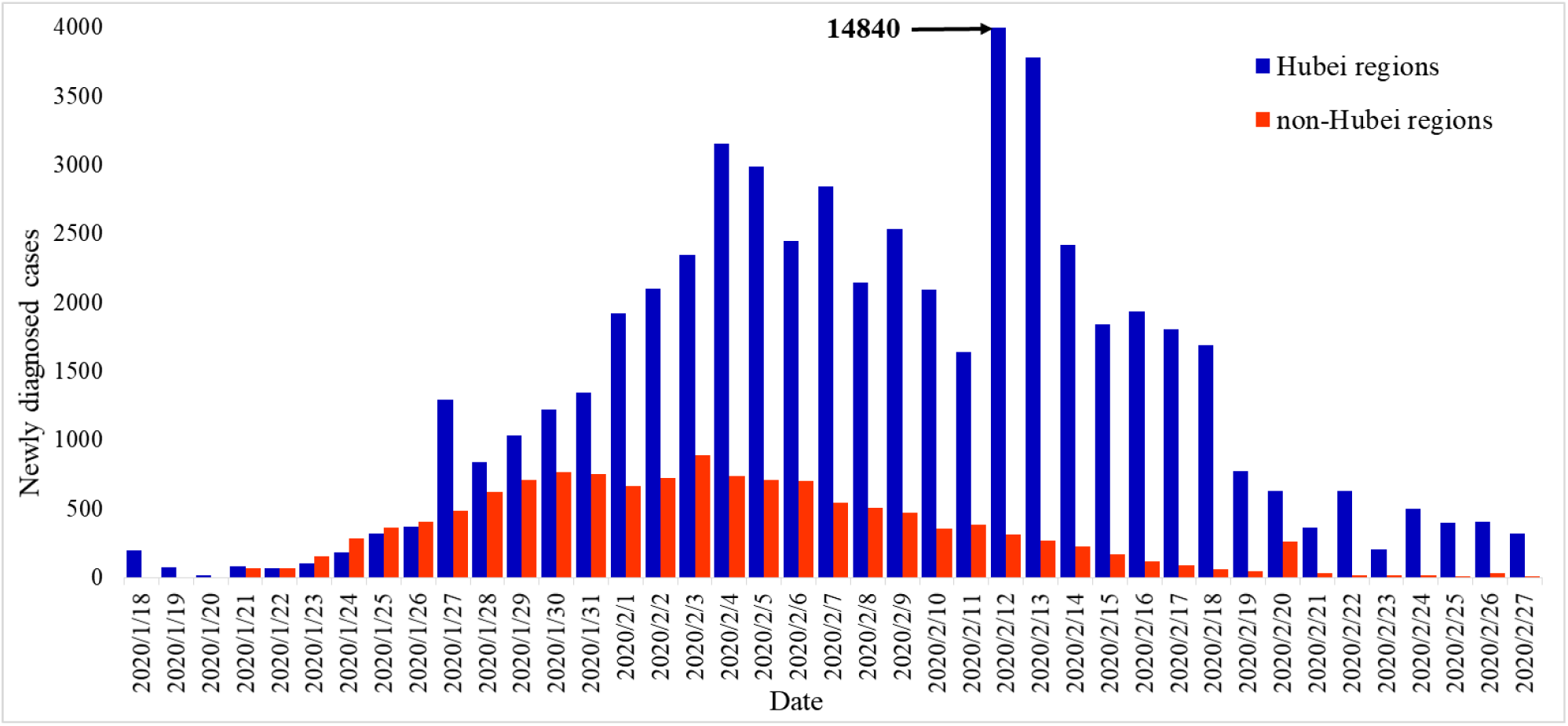
The number of newly diagnosed cases in Hubei and non-Hubei regions in China Mainland

The SARS-CoV-2 epidemic is still rapidly growing and spread to more than 42 countries as of February 27, 2020. At present, the most serious countries outside China are South Korea, Italy, Iran, and Japan. It is worrying that although some measures have been taken, the current prevention and control measures of these countries may still be insufficient. None of them has reached the level of prevention and control in China’s moderately affecting Provinces some time ago. The controllable window period is narrowing, the international community should learn from lessons of Wuhan and experience from China. It’s Time for Action.

## Data Availability

The data are from Baidu Migration (http://qianxi.baidu.com) and the National Health Commission of China (http://www.nhc.gov.cn/xcs/yqtb/list_gzbd.shtml).

## Author Contributions

Y. X. collected and pre-processed the data, Z.Y., Y.C. and Y. X. analysed the data, Z.Y., Y. X., Z. D., J. H. and Y.C. wrote the paper. All the authors reviewed the manuscript.

This work was supported by the Scientific Research Foundation of Education Office of Hunan Province, China [17A096]; the National Natural Science Foundation of China [61701177].

